# Knowledge and practice of road safety measures among motor-bikers in Bangladesh: a cross-sectional study

**DOI:** 10.1101/2023.07.25.23293162

**Authors:** Sadhan Kumar Das, Tahazid Tamannur, Arifatun Nesa, Abdullah Al Noman, Piue Dey, Shuvojit Kumar Kundu, Hafiza Sultana, Baizid Khoorshid Riaz, ANM Shamsul Islam, Golam Sharower, Bablu Kumar Dhar, Mohammad Meshbahur Rahman

**Author notes:** Correspondence to: Mohammad Meshbahur Rahman, Assistant Professor, Department of Biostatistics, National Institute of Preventive and Social Medicine (NIPSOM), Dhaka 1212, Bangladesh.

## Abstract

**Introduction:** Road traffic accidents (RTAs) including motorbike accidents are the leading cause of death for teenagers between 15 and 29 and have been a serious concern in low- and middle-income countries like Bangladesh. Therefore, this study aims to assess the level of knowledge and practice on road safety measures among motorbikers in Bangladesh.

**Methods:** This cross-sectional study was conducted from January 2022 to December 2022 among 350 motor-bikers of Dhaka, the capital of Bangladesh via a series of face-to-face interviews. Motor bikers who regularly ride a motorcycle were interviewed about their road rafety knowledge and practices through two-stage cluster sampling technique. Frequency distribution, independent sample t-test, and One-way ANOVA were performed.

**Results:** Out of the 350 motorbikers, only 54.6% had good knowledge and 16.9% had poor knowledge on the signs and safety regulations of roads. Moreover, only 50.6% of respondents followed good practices while 23.4% followed poor practices of road safety measures. One way ANOVA analysis demonstrates that the average knowledge score was significantly (p<0.05) higher among higher-educated, unmarried and non-smokers. Additionally, higher education level, non-smoking status and being Muslim were significantly (P<0.05) associated with good road safety practices.

**Conclusions:** The overall good knowledge level and practices of road safety measures among the motorbikers were not satisfactory albeit the majority of them knew individual signs and regulations. Therefore, this study suggests that education and strict enforcement of traffic rules may increase their knowledge and practice behavior regarding road safety which in turn would minimize traffic injuries and fatalities.

## Introduction

Globally, road traffic accidents (RTA) pose a serious threat to public health and are the leading cause of death among children, adolescents and young adults. Each year, around 1.3 million people die and about 20–50 million people suffer from traffic-related injuries that cost $518 billion worldwide [1,2]. Moreover, it is predicted to be the 5^th^ leading cause of death [3] and 3^rd^ leading cause of lost disability-adjusted life years by 2030 [4]. Worriedly, more than 90% of the fatalities occurred in low- and middle-income countries [2,5]. Studies estimated that the traffic fatality rate in low- and middle-income countries is 20.2 deaths per 100,000 people, which is much higher than the rate in high-income countries (12.6 deaths per 100,000 people) [6]. According to a global assessment, around 46% of all traffic fatalities involve pedestrians, cyclists, and riders of motorized two-wheelers, as well as their passengers [7]. Road traffic injuries place a huge strain on healthcare services in terms of financial resources, bed occupancy and demand placed on health professionals [8]. The World Health Organization (WHO) reported that Asia is the continent with the highest (6.5%) rate of motorcycle fatalities per 10,000 bikes [9]. Six Asian nations such as Bangladesh, Cambodia, Laos, Thailand, India, and Myanmar have fatality rates that are higher than the average RTAs fatality rate of Asia.

From the last few couple of years, Bangladesh has been facing numerous public health challenges including the outbreak of viral diseases (e.g., COVID-19, Dengue), child and elderly nutritional needs, and healthcare management [10–17]. Along with this, the country has been noted for excessive RTA over the previous five years [18]. According to the Passenger Welfare Association of Bangladesh reports, a non-government authority, more than 6000 people died each year for the last eight years due to road accidents in Bangladesh which also injured over 100000 individuals [19]. However, this is not the clear picture since under-reporting is very common in countries like Bangladesh where strong road traffic system is scarce. Surprisingly, the Bangladesh Road Traffic Authority (BRTA) started to publish reports on road accidents by themselves this year. Before that, only FIR report from police was counted as official cases [20]. As a result, this impedes the Sustainable Development Goal’s target to reduce road traffic injuries and fatalities by half by 2030.

Similar to global road accident cases, motorcycle accidents and fatalities have also been increasing in Bangladesh due to a dramatic rise in motorbike ownership. Motorcycles consisted of approximately 70% of all registered motor vehicles in the country with around 3.5 million [21,22]. However, the authority issued only 22 million licenses which means a large number of unlicensed vehicles run on the road and this is direct evidence for a high rise in RTA by motorcycles. The most alarming fact is that Bangladesh is the country with the highest motorcycle death rate, 28 per 10,000 motorcycles [23].

Road accidents are multi-cause events that frequently result from many factors, including driver error, the state of the road, and the condition of the vehicle [24–26]. It entails significant financial consequences to fatalities, injuries, and lost potential revenue, as well as significant human suffering [27,28]. Methods and techniques for minimizing the risk of a person using the road network, being killed or seriously injured are referred to as road traffic safety [29]. The best practice of road safety strategies emphasizes the preclusion of major injury and death crashes despite human fallibility [30]. However, if drivers do not understand the information on safe driving behavior that is encoded in the traffic signs, the signs will not be able to successfully perform their intended goals [31,32]. Studies on how drivers interpret traffic psychological and demographic factors are still alarming [32–34].

As previously described, motorbikes are the prevalent vehicles in Bangladesh and RTA is the predominant cause of death for individuals aged 15-29 years, the purpose of this study was to assess the level of knowledge and practice of road safety measures among motorbikers in Bangladesh. To the best of our knowledge, little research has been done on road traffic accidents in Bangladesh in general [28,35–37], and no study has been conducted on motorbikers. Therefore, the findings of this study would be helpful for motor-bikers in increasing their knowledge and practice level regarding road safety. Moreover, it would be evidence for assisting the policy leaders to reappraise the policies regarding road safety and to devise appropriate action-based policies.

## Methods

### Ethical consent and permission for data collection

This study was approved by the institutional review board of the National Institute of Preventive and Social Medicine (NIPSOM), Bangladesh (Ref No: NIPSOM/IRB/2017/09). Both writtena and verbal consent were taken before inititing the interview. A brief of the aims and objectives was given to the participants. Participants who agreed to give consent were finally included in the study.

### Study setting and participants

This study was a cross-sectional study conducted by covering in all the administrative wards of North and South City Corporation, Dhaka from 1st January to 31st December 2022.

### Sampling technique and sample size

Two-stage cluster sampling technique was applied in this study. In the first stage, all the major traffic points in Dhaka North and Dhaka South city corporation were listed and considered as clusters. In the second stage, twenty traffic points were randomly selected and motor-bikers who gave consent were included in the study. The sample size of the study was calculated by using the formula below:

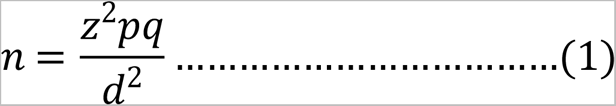

Here, n= assumed/ desired sample size; z= the standard normal deviation; usually set at 1.96 at a 95% confidence level; p= percentage of practice is 17.6% = 0.176, obtained by the literature search [38]; q= 1-p; and d= Margin of error (5%) = 0.05.

Using equation (1), the sample size for the study when p= 0.176 is

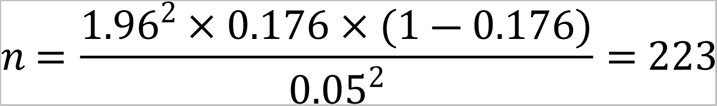

Initially, the study considered 223 participants as the required sample size. However, since no study was found that studied with a sufficient sample size, we additionally considered a usable 1.5 design effect and a 5% non-response rate. Finally, the sample size was 350.

### Selection criteria

The inclusion criteria were: (i) Motor bikers who regularly ride a motorcycle; (ii) male and aged 18 or above.

### Outcome measures

Respondents’ socio-demographic variables such as age, educational status and monthly income and their habits such as smoking behaviors and alcohol consumption patterns were considered independent variables. On the other hand, motorbikers’ knowledge and practice regarding road safety measures were dependent variables. These are presented in supplementary Table S1.

### Data collection tool and procedures

A structured questionnaire was used to collect data from the participants. Thirteen traffic signs provided by the BRTA and commonly used in the country were applied to assess respondents’ knowledge. We used 22 variables to assess bikers’ knowledge and 16 variables to assess their practice score regarding road safety measures. The average scoring technique was used in computation and their descriptive statistics including percentiles were observed. According to the percentile approach, the knowledge and practices were classified into three levels-poor (<25% percentile, score: <0.727); average (25%-49% percentile, score: 0.727-0.817); and good (≥50% percentile, score: ≥0.818) [39].

### Statistical analysis

To ensure the reliability and validity of the study results, we used Cronbach’s alpha. The reliability coefficient values for the variables related to knowledge and practice were found 0.50 and 0.26). Descriptive statistics were performed to present the socio-demographic characteristics and mean knowledge and practice scores of participants. Since both knowledge and practice scores didn’t follow normality, we performed an independent sample t-test and one-way ANOVA test to show the mean knowledge and practice difference between two (e.g.: rural versus urban) and more than two (e.g.: different age groups) groups respectively. A p-value of 5% was considered significant at the 95% Confidence Interval (CI). All the analysis was performed in Microsoft Office-2019 and SPSS (Version-26) software.

## Results

### Socio-demographic characteristics of the respondents

Table 1 shows the socio-demographic characteristics of the respondents. It was found that the majority of the respondents (42.86%) were aged between 30 and 39 years followed by respondents aged 18 to 29 years. Only 36.3% had completed tertiary education while 52% had secondary or higher secondary education. Almost half of the participants had a monthly family income of 20000-30000 BDT. Moreover, the majority of them were engaged (67.7%), followed by service. Urban participants were more prevalent since the study covered participants from the capital city. While alcohol consumption was at the least (only 1%), the prevalence of smoking was high (61.7%) among participants.

**Table 1.**
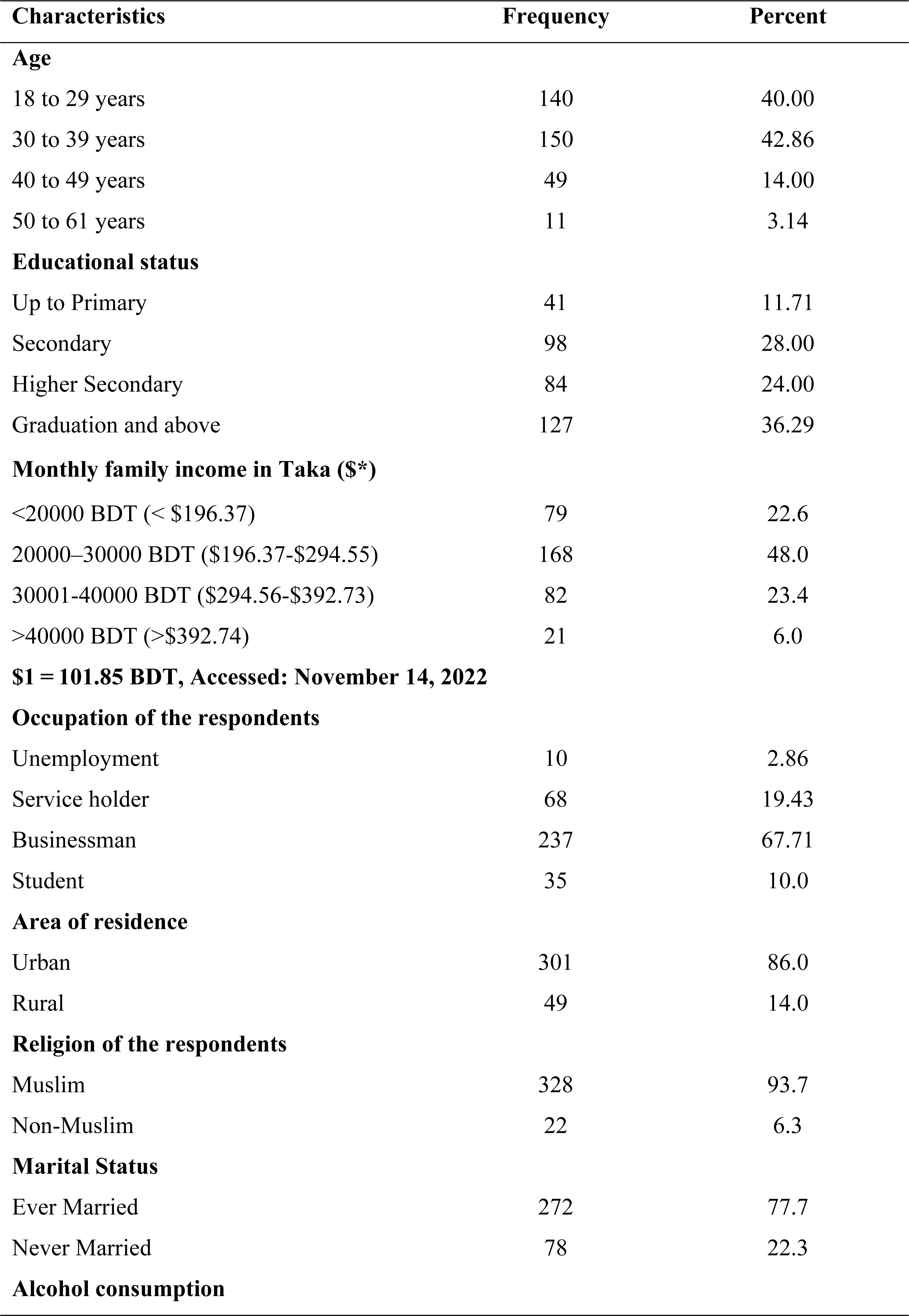
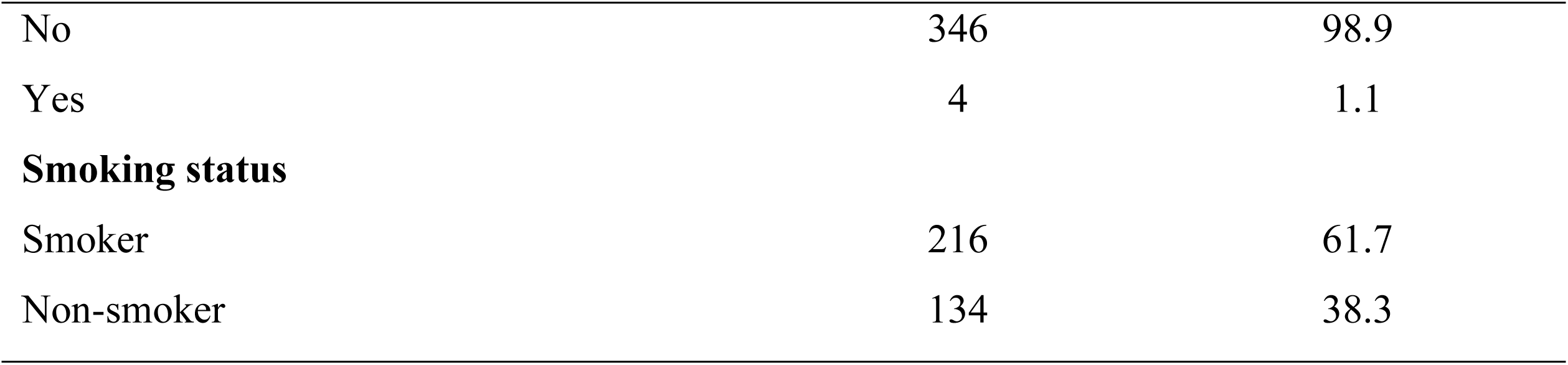
Socio-demographic characteristics of the motor-bikers in Dhaka city.

### Knowledge regarding road safety measures among respondents

Table 2 represents participants’ knowledge about road safety measures. Among the 350 respondents, more than 80% had insurance while only 56.3% of them knew the maximum speed limit for driving in Dhaka city. Surprisingly, the ‘No stopping’ sign was unknown to about 80% of motorbikers, while about half of them did not know the ’Stopping’ sign. However, the majority of the respondents knew about the ‘No horn’ (98.6%), ‘U-turn’ (97.1%), ‘Zebra-crossing’ (94.3%) and ‘Speed breaker sign (85.4%). Moreover, almost all of them knew wearing a helmet and updating their documents is mandatory and drinking alcohol (98.3%), using mobile phones (98.3%), playing music ((96.6%), frequent overtaking (96.9%), talking to other (92.3%) while driving is dangerous and/or prohibited. (Table 2).

**Table 2.**
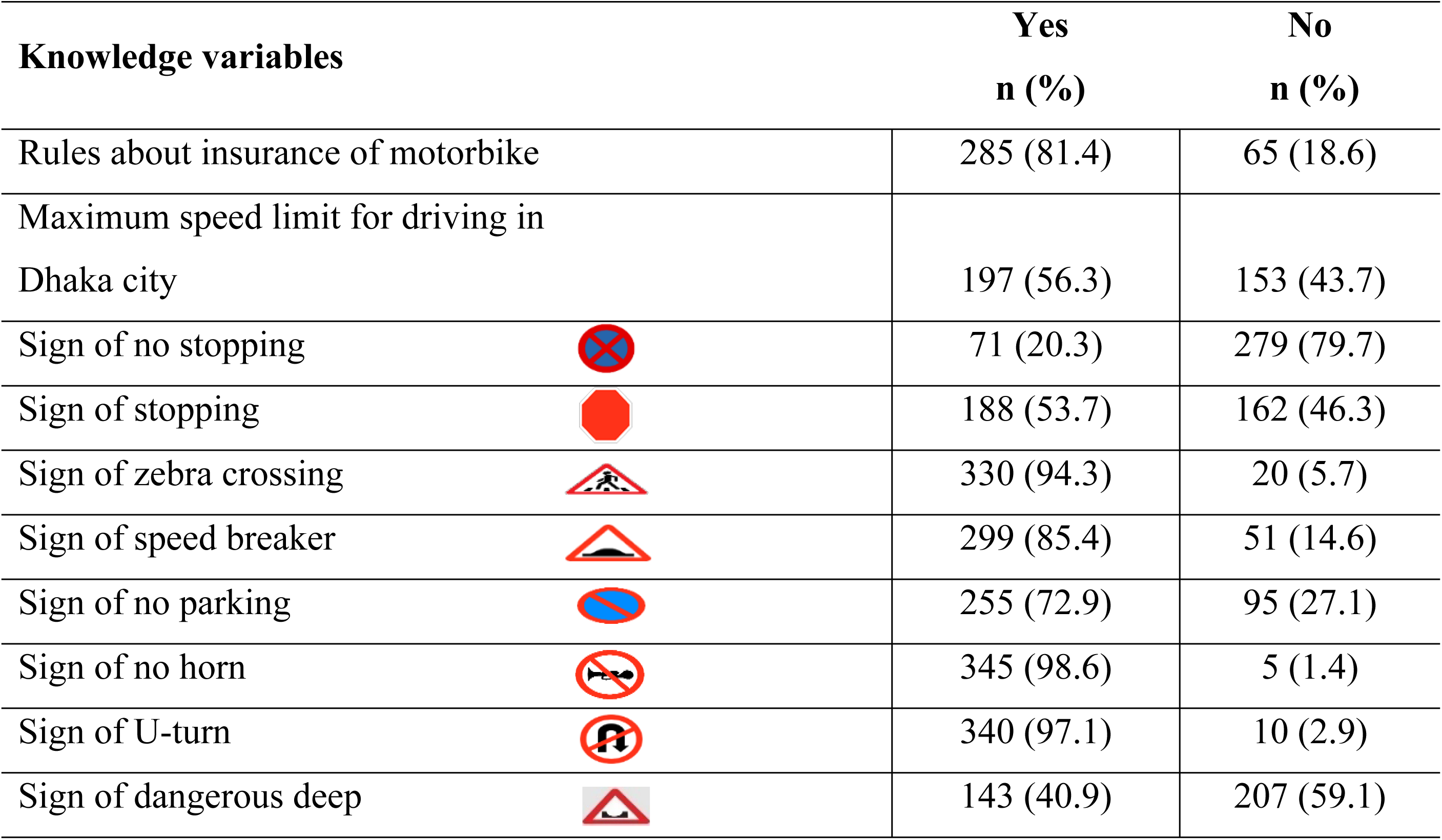
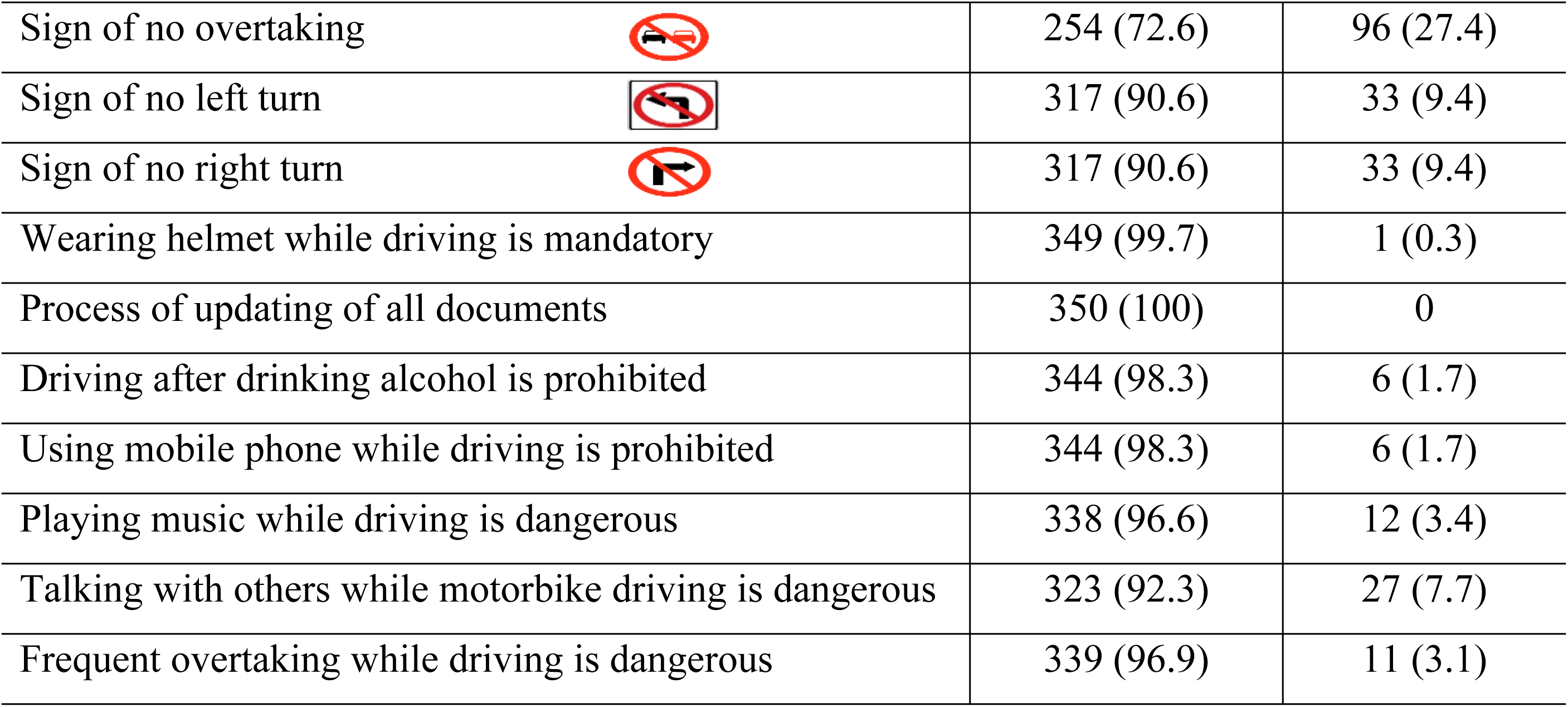
Knowledge about road safety measures among respondents.

| **Knowledge variables** |  | **Yes**<br>**n (%)** | **No**<br>**n (%)** |
| --- | --- | --- | --- |
| Rules about insurance of motorbike |  | 285 (81.4) | 65 (18.6) |
| Maximum speed limit for driving in Dhaka city |  | 197 (56.3) | 153 (43.7) |
| Sign of no stopping |  | 71 (20.3) | 279 (79.7) |
| Sign of stopping |  | 188 (53.7) | 162 (46.3) |
| Sign of zebra crossing |  | 330 (94.3) | 20 (5.7) |
| Sign of speed breaker |  | 299 (85.4) | 51 (14.6) |
| Sign of no parking |  | 255 (72.9) | 95 (27.1) |
| Sign of no horn |  | 345 (98.6) | 5 (1.4) |
| Sign of U-turn |  | 340 (97.1) | 10 (2.9) |
| Sign of dangerous deep |  | 143 (40.9) | 207 (59.1) |
| Sign of no overtaking |  | 254 (72.6) | 96 (27.4) |
| Sign of no left turn |  | 317 (90.6) | 33 (9.4) |
| Sign of no right turn |  | 317 (90.6) | 33 (9.4) |
| Wearing helmet while driving is mandatory |  | 349 (99.7) | 1 (0.3) |
| Process of updating of all documents |  | 350 (100) | 0 |
| Driving after drinking alcohol is prohibited |  | 344 (98.3) | 6 (1.7) |
| Using mobile phone while driving is prohibited |  | 344 (98.3) | 6 (1.7) |
| Playing music while driving is dangerous |  | 338 (96.6) | 12 (3.4) |
| Talking with others while motorbike driving is dangerous |  | 323 (92.3) | 27 (7.7) |
| Frequent overtaking while driving is dangerous |  | 339 (96.9) | 11 (3.1) |

### Practice regarding road safety among respondents

Regarding good practices, the majority of the participants answered affirmatively regarding having insurance for their bikes (77.7%) and updating their licensing documents regularly (99.1%). Moreover, most of them followed road safety practices such as using a helmet (99.7%) and following the road speed limit while driving (91.4%) and being patient while pedestrians take time while crossing roads (96.0%). On the other hand, the majority avoided bad driving habits such as driving while drunk, using mobile phones and talking with others while driving. However, 88.3% of respondents practiced overtaking frequently while driving and over 60% were penalized by police for violating traffic laws (Table 3).

**Table 3.**
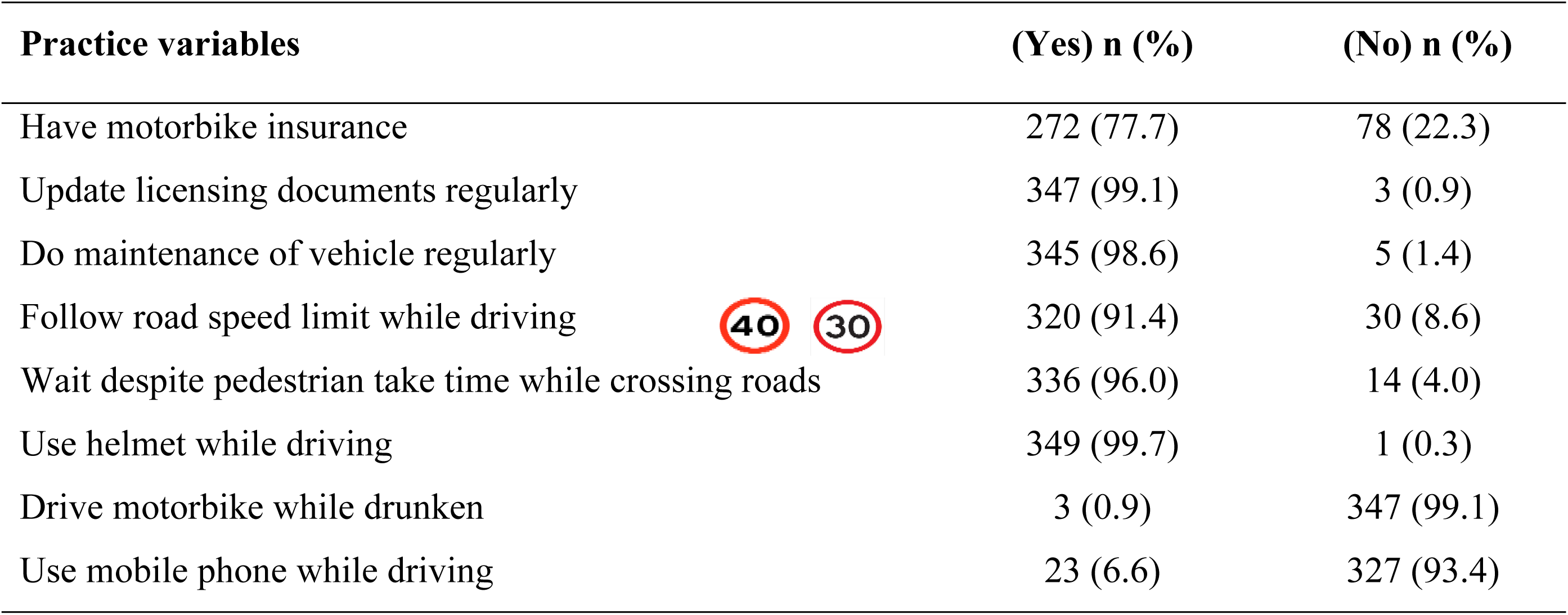
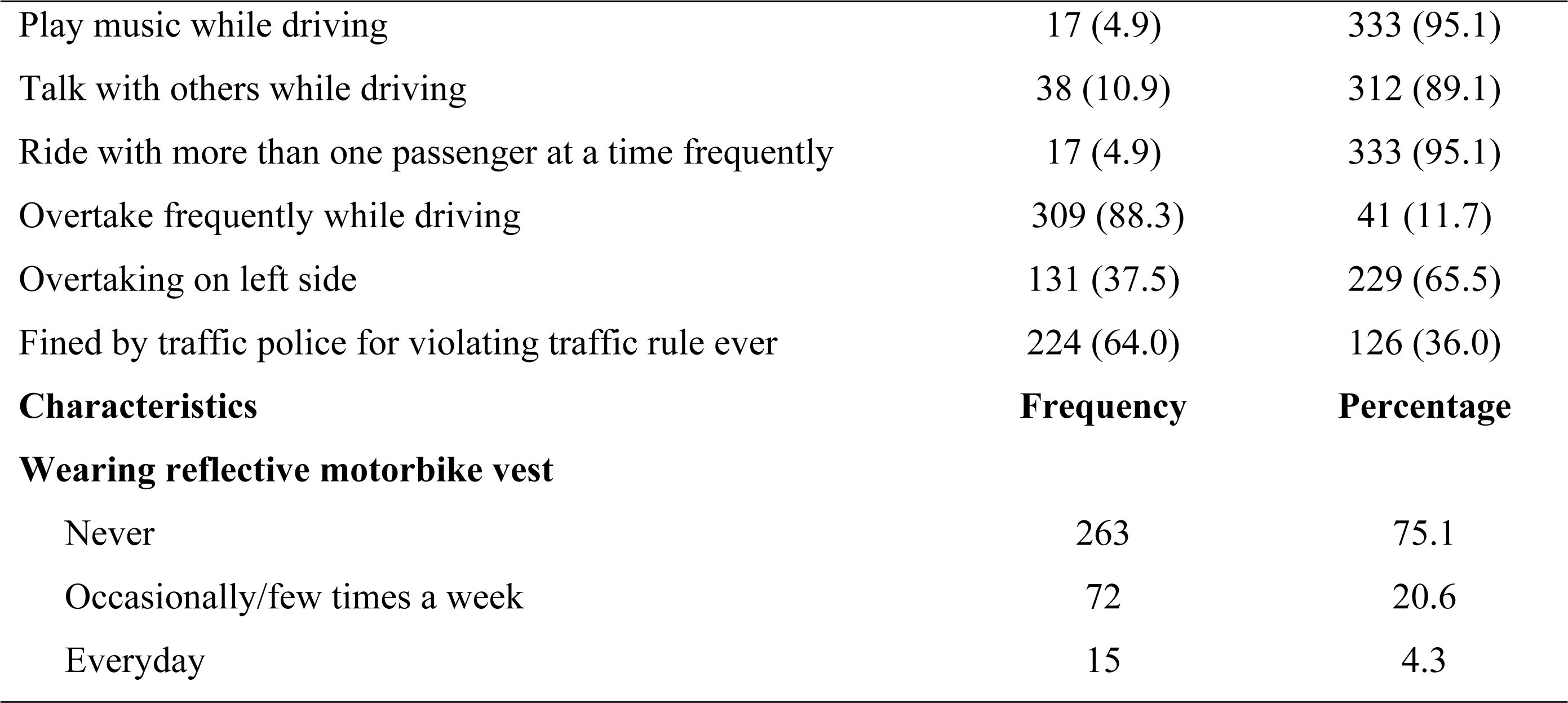
Distribution of practice of road safety measures among motor-bikers.

| Practice variables | (Yes) n (%) | (No) n (%) |
| --- | --- | --- |
| Have motorbike insurance | 272 (77.7) | 78 (22.3) |
| Update licensing documents regularly | 347 (99.1) | 3 (0.9) |
| Do maintenance of vehicle regularly | 345 (98.6) | 5 (1.4) |
| Follow road speed limit while driving | 320 (91.4) | 30 (8.6) |
| Wait despite pedestrian take time while crossing roads | 336 (96.0) | 14 (4.0) |
| Use helmet while driving | 349 (99.7) | 1 (0.3) |
| Drive motorbike while drunken | 3 (0.9) | 347 (99.1) |
| Use mobile phone while driving | 23 (6.6) | 327 (93.4) |
| Play music while driving | 17 (4.9) | 333 (95.1) |
| Talk with others while driving | 38 (10.9) | 312 (89.1) |
| Ride with more than one passenger at a time frequently | 17 (4.9) | 333 (95.1) |
| Overtake frequently while driving | 309 (88.3) | 41 (11.7) |
| Overtaking on left side | 131 (37.5) | 229 (65.5) |
| Fined by traffic police for violating traffic rule ever | 224 (64.0) | 126 (36.0) |
| **Characteristics** | **Frequency** | **Percentage** |
| **Wearing reflective motorbike vest** |  |  |
| Never | 263 | 75.1 |
| Occasionally/few times a week | 72 | 20.6 |
| Everyday | 15 | 4.3 |

### Overall knowledge and practice level of the respondents

Fig 1 depicts the level of average knowledge and practices of respondents and their association with educational status. It can be seen that only 54.5% had good knowledge, while 16.9% had poor knowledge (Fig 1a). Similarly, only half of the respondents (50.6%) had a good practice level while 23.4% had a poor practice level (Fig 1b).

**Fig 1.**
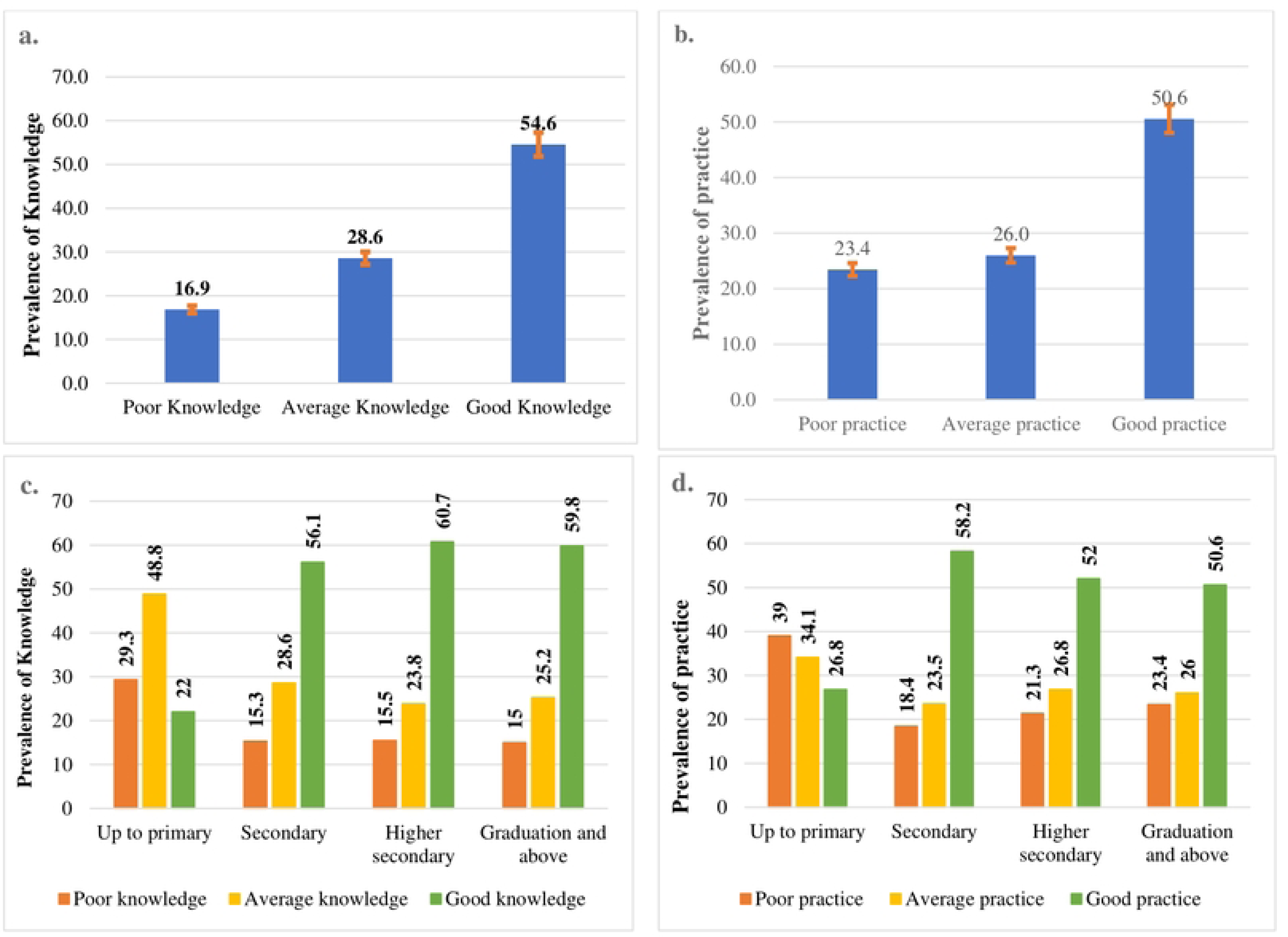
(a) Distribution of overall knowledge level; (b) Distribution of overall practices level; c) Association of knowledge with respondents’ educational status; (d) Association of practice with respondents’ educational status.

Higher knowledge levels and following good practices were associated with higher educational status (Fig 1c and Fig 1d). Approximately, over 55% of participants with secondary, higher secondary and tertiary education had good knowledge of road safety, while the majority of respondents with primary education had poor to average knowledge (Fig 1c). Similarly, more than 50% of motorbikers who completed secondary, higher secondary and tertiary education followed good practices while most of the motorbikers with primary education did not follow good practices (Fig 1d).

### Variation in Knowledge and Practices of the respondents

A significant difference in respondents’ knowledge and practice with socio-demographic characteristics was observed (Table 4). Results found that the average knowledge score was significantly higher among younger adults aged 18-29 years than that among the older population aged 50 and above [mean knowledge score: 0.80 (0.79-0.82) versus 0.75 (0.69-0.82); p=0.04]. Similarly, the mean knowledge score was higher among participants with secondary, higher secondary and tertiary education compared to the primary education group. Moreover, knowledge score was higher among non-married [0.81 (0.79-0.83) versus 0.78 (0.77-0.79); p=0.006] and non-smokers [0.80 (0.79-0.82) versus 0.78 (0.77-0.79)]; p=0.026] than their counterparts (Table 4).

**Table 4.** Independent sample t-test and one-way ANOVA test for knowledge and practice score.

| Characteristics | n | Mean knowledge<br>score (C.I) | P*<br>value | Mean practice<br>score (C.I) | P# value |
| --- | --- | --- | --- | --- | --- |
| Age (years) |  |  |  |  |  |
| 18 to 29 | 140 | 0.80(0.79-0.82) | 0.043 | 0.78 (0.76-0.79) | 0.810 |
| 30 to 39 | 150 | 0.79(0.78-0.81) |  | 0.77(0.76-0.79) |  |
| 40 to 49 | 49 | 0.76(0.73-0.79) |  | 0.79(0.76-0.81) |  |
| 50 to 61 | 11 | 0.75(0.69-0.82) |  | 0.77(0.71-0.84) |  |
| Educational status |  |  |  |  |  |
| Up to Primary | 41 | 0.73(0.70-0.76) | 0.0001 | 0.74(0.71-0.77) | 0.011 |
| Secondary | 98 | 0.79(0.77-0.81) |  | 0.79(0.77-0.81) |  |
| Higher Secondary | 84 | 0.81(0.79-0.83) |  | 0.77(0.75-0.79) |  |
| Graduation & above | 127 | 0.79(0.78-0.81) |  | 0.78(0.77-0.79) |  |
| Occupation |  |  |  |  |  |
| Unemployed | 10 | 0.77(0.72-0.83) |  | 0.74(0.69-0.79) |  |
| Service holder | 68 | 0.79(0.77-0.81) | 0.620 | 0.79(0.77-0.81) | 0.491 |
| Businessman | 237 | 0.79(0.78-0.80) |  | 0.78(0.77-0.81) |  |
| Student | 35 | 0.81(0.78-0.83) |  | 0.77(0.77-0.79) |  |
| **Monthly family income in Taka (\$\*)** | | | | | |
| <20000 BDT (< \$196.37) | 79 | 0.79(0.77-0.81) | | 0.787(0.77-0.81) | |
| 20000–30000BDT (\$196.37-\$294.55) | 168 | 0.80(0.79-0.81) | 0.149 | 0.77(0.76-0.79) | 0.514 |
| 30001-40000 BDT (\$294.56-\$392.73) | 82 | 0.77(0.75-0.79) | | 0.76(0.75-0.79) | |
| >40000 BDT (>\$392.74) | 21 | 0.77(0.72-0.83) | | 0.78(0.77-0.79) | |
| **Dollars, considering \$1 = 101.85 BDT, Accessed: November 14, 2022** | | | | | |
| **Residents** |  |  |  |  |  |
| Urban | 301 | 0.78(0.78-0.80) | 0.927 | 0.77(0.76-0.78) | 0.128 |
| Rural | 49 | 0.79(0.76-0.82) |  | 0.79(0.79-0.81) |  |
| **Religion** |  |  |  |  |  |
| Muslim | 328 | 0.79(0.78-0.79) | 0.168 | 0.77(0.77-0.78) | **0.025** |
| Non-Muslim | 22 | 0.81(0.78-0.84) |  | 0.73(0.69-0.77) |  |
| **Marital status** |  |  |  |  |  |
| Ever Married | 272 | 0.78(0.77-0.79) | **0.006** | 0.78(0.77-0.79) | 0.470 |
| Never Married | 78 | 0.81(0.79-0.83) |  | 0.77(0.75-0.79) |  |
| **Alcohol status** |  |  |  |  |  |
| No | 346 | 0.79(0.78-0.79) | 0.867 | 0.78(0.77-0.79) | 0.675 |
| Yes | 4 | 0.79(0.70-0.89) |  | 0.75(0.56-0.94) |  |
| **Smoking status** |  |  |  |  |  |
| Smoker | 216 | 0.78(0.77-0.79) | **0.026** | 0.77(0.76-0.78) | **0.008** |
| Non-smoker | 134 | 0.80(0.79-0.82) |  | 0.79(0.78-0.81) |  |
| *t-test/ANOVA test for knowledge; # t-test/ANOVA test for practice |  |  |  |  |  |

Regarding practices, the mean practice score was associated with educational status, religion and smoking status. Similar to knowledge level, the average practice score was higher among participants with secondary, higher secondary and tertiary education compared to the primary group. Additionally, average practice score was significantly higher [mean practice score: among Muslims [0.77 (0.77-0.78) versus 0.73 (0.69-0.77); p=0.025].

## Discussion

In Bangladesh, a significant increase in motorbike accidents created an unnecessary healthcare burden that required urgent attention. Our study highlighted that despite increasing RTA, motorbikers lack adequate knowledge and are associated with poor road traffic practices. However, an increase in knowledge level was observed in young adults, higher education level, non-smokers and non-married respondents. Similarly, good practices were associated with education level and smoking status.

Among our participants, the majority had insurance. Regrettably, only 56.3% of them knew the maximum speed limit for driving in Dhaka city while it has been observed that the highest number of motorbike and bike accidents occurs in Dhaka division (30). Most of the respondents could recognize most of the signs such as ‘stopping’, ‘zebra crossing’, ‘u turn’ and ‘no overtaking’. However, only 20% of them could recognize the sign of ‘no stopping’ This finding is comparable to studies in other South Asian countries. A study in India reported that among college students (aged 17-23), only 65.1% correctly identified the road signs while another study among college students in Pakistan showed that 63.6% correctly identified the signs [40] In a study by Reang and Tripura, it was found that nearly 90% of respondents did not know about the ‘no stopping’ sign [41]. Interestingly, the majority of motorbikers knew that wearing a helmet while driving is mandatory. Different studies found different results regarding wearing helmets, albeit nearly all countries mandate this law. For instance, a study in India by Jothula and Sreeharshika revealed that around 98% of respondents knew the law [27], while another study by Siviroj et al found that around 56% of motorbikers knew about the potential danger of not wearing a helmet [42]. Similarly, over 90% of our study participants were aware that driving after drinking alcohol or using a mobile phone while driving is prohibited. Moreover, most of them knew that playing music, talking with others and overtaking frequently during riding are dangerous behaviors that could kill them. Compared to many other studies done in developing or least developed countries, respondents of this study were more aware of the laws. Jothula and Sreeharshika found that only 6.8% of the respondents of a study in India knew the permissible limit of alcohol [43]. Similarly, Tajvar et al. depicts that participants of their study believed that eating (49.8% agree) and drinking during driving are dangerous (34.9% agree) [44].

In accordance with the knowledge level, the majority of our participants followed good practices. The majority of the motorbikers wore helmets (99.7%) and followed the average speed limit (91.4%). Moreover, a small percentage of motorbikers reported driving after drinking alcohol (0.3%), using their phones (6.6%), playing music (4.6%) or talking with others (10.9%) while driving. Conversely, many studies found a lower percentage of respondents wearing helmets. Jennissen *et al*. found that 64% never wear helmets while driving [45]. Furthermore, Wadhwaniya found that only 44.5% [46] and Jothula and Sreeharshika found that only 25.9% of respondents wear helmets while driving [47]. Helmets are recommended to prevent severe injuries and wearing helmets for motorbikers is a law in most countries. Studies found that it can minimize the risk of death to 42%-69% [48]. However, 88.3% of our participants reported overtaking frequently, albeit most of them knew it is dangerous. In comparison, only 44.4% of participants in an Iranian study performed overtaking practice When it comes to young people, overtaking is one of the most dangerous [44] maneuvers they display frequently which caused a large number of injuries and fatalities. A German study depicts that around 6 percent of the accidents of which around 9 percent of deaths occurred in 2014 are due to overtaking practices [49]. In addition to these, most of our participants (64.0%) were penalized by police at least once due to traffic violations which indicate their poor practice habits.

However, although the majority of our respondents could identify signs individually or reported being aware of some rules and regulations, their overall knowledge level of road safety was not satisfactory. Only 54.5% had good knowledge while the prevalence of poor knowledge was also noticeable. Likewise, as expected, only half of them followed good practices and a significant number of participants followed poor practices. Another similar study conducted by Baniya and Timilsina (2018) in Nepal showed that more than half (59.2%) of the respondents had moderately adequate knowledge [39]. An interesting finding was that both knowledge and good practice levels were associated with respondents’ educational status.

Some socio-demographic factors were associated with both knowledge level and practices of road safety measures. One-way ANOVA test revealed that knowledge score differs significantly among respondents with different educational levels, marital and smoking status. The knowledge level of participants with a higher secondary degree was significantly higher than people with other degrees. Although most of the world’s studies reported that the majority of road accidents and death involve young people aged between 20 and 29 years [50]. Moreover, people who were married at any time and non-smokers tend to have higher knowledge scores. Although no studies reported these factors as contributing factors to increasing knowledge level, it is perceivable that people who are married and non-smokers tend to be more aware and conscious regarding life. Education levels were also found to be a contributing factor in higher practice scores as participants with higher secondary degrees had a significantly higher score than people with other degrees. Moreover, significantly higher practice scores were observed in Muslims and non-smokers. A study by Bachani et al. (2017) found that educated persons have a lower chance of being smokers than uneducated ones. As the majority of the respondents included in the study were educated, thus majority were non-smokers and had good knowledge and followed good practices on road safety measures [51].

### Recommendations and future directions

Road traffic injuries have become one of the major health concerns worldwide which constituted about 11% of the global burden of disease in 2015 [52]. According to the WHO, the annual mortality rate due to road accidents per capita in Bangladesh is twice the average rate in developed countries and is the highest in South Asia [53]. The government is lagging far behind in achieving the target of the Sustainable Development Goals of reducing the number of injuries and deaths by half by 2030. Hence, urgent attention is needed to tackle this silent public health threat.

#### Training for drivers

Bangladesh Road Transport Authority, the sole authority providing driving licenses should arrange training or educational campaigns on road safety measures. Currently, they provide training on only driving and upgrading vehicles. Many countries introduced ‘Road Safety Education’ programs that considerably reduced morbidity and mortality [54]. However, it was also observed that the effects of many of these interventions did not sustain for a long time [55] and hence, training or campaigns need to be arranged quarterly or half-yearly.

#### Minimizing under-reporting

Under-reporting is another concern for least-developed countries like Bangladesh that prevent people from perceiving the true magnitude of road accidents. While the authority estimated annual road accident-related deaths in 2015 was 2,538, the WHO estimated the number may range from 20,736 and 21,316 [56]. Therefore, the authority should take every case into account and publish true reports publicly that would facilitate mass awareness.

#### Develop a robust referral system

Like many middle and low-income countries, Bangladesh lacks a robust referral system that includes health risk and hazard assessment and direct medical costs. Severe injuries could lead to death when urgent treatment is not available. A comprehensive referral system comprises differentiating severe cases, timely transportation, prompt communication between facilities and immediate care at treatment facilities. Policymakers, public health experts, nongovernmental advisory organizations (Nirapad Sarak Chai, Road Safety Foundation) and leaders of vehicle owners’ associations should work collaboratively to devise an effective scheme.

#### Stringent regulations and awareness

The government could initiate road safety education at school and college levels since road accidents are more common among adolescents and teenagers. All drivers should be licensed and their data should be stored with the authority so that nobody can avoid punishment if involved with a major accident. Finally, it is more of a self-responsibility to comply with good safety practices.

### Conclusion

The overall knowledge and practice behavior of motorbikers was not satisfactory. Higher education and quiting smoking during the drive were the significant factor that contributed to increase knowledge and good road safety practice. The findings of the study are important for the authority and policymakers of Bangladesh to develop pragmatic road safety contrivances that would considerably reduce morbidity and mortality, and help achieve targets of sustainable development goals.

## Acknowledgements

We acknowledge the department of Biostatistics for their technical support duering the study. We are also grateful to all perticipants included this study.

## Authors contributions

**Conceptualizations-** Mohammad Meshbahur Rahman, Hafiza Sultana and Sadhan Kumar Das

**Formal analysis-** Sadhan Kumar Das and Mohammad Meshbahur Rahman

**Investigation-** Sadhan Kumar Das, Tahazid Tamannur, Arifatun Nesa and Mohammad Meshbahur Rahman

**Methodology-** Sadhan Kumar Das and Mohammad Meshbahur Rahman

**Project administration-** Sadhan Kumar Das, Abdullah Al Noman, Shuvojit Kumar Kundu, Hafiza Sultana, Baizid Khoorshid Riaz, ANM Shamsul Islam, Golam Sharower and Mohammad Meshbahur Rahman

**Supervision-** Mohammad Meshbahur Rahman

**Validation-** Hafiza Sultana, Baizid Khoorshid Riaz, ANM Shamsul Islam, Golam Sharower, Bablu Kumar Dhar and Mohammad Meshbahur Rahman

**Visualization-** Sadhan Kumar Das, Abdullah Al Noman, Shuvojit Kumar Kundu and Mohammad Meshbahur Rahman

**Writing-original draft-** Sadhan Kumar Das, Tahazid Tamannur, Arifatun Nesa, Abdullah Al Noman, Shuvojit Kumar Kundu, Bablu Kumar Dhar and Mohammad Meshbahur Rahman

**Writing -review and editing** Sadhan Kumar Das, Tahazid Tamannur, Arifatun Nesa, Abdullah Al Noman, Shuvojit Kumar Kundu, Hafiza Sultana, Baizid Khoorshid Riaz, ANM Shamsul Islam, Golam Sharower, Bablu Kumar Dhar and Mohammad Meshbahur Rahman

## Data availability statement

The datasets generated and/or analyzed during the current study are available from the corresponding author on reasonable request to.

